# Dosage individualization proposed for anti-gout medications among the patients with gout: a bicentric study in Nepal

**DOI:** 10.1101/2020.12.08.20245860

**Authors:** Binaya Sapkota, Suraj Chaudhary, Prakash Gurung, Anisha Humagain, Sujan Sapkota

## Abstract

**Background:** The conventional one-size-fits-all approach has been criticized for almost all drugs used especially for chronic diseases, including gout. The present study was aimed to individualize and optimize the dose of anti-gout medications among gout patients.

**Methods and findings:** Bicentric cross-sectional study was carried out among 384 randomly selected new gout patients visiting two gout treatment centers at Lalitpur Metropolitan City, Nepal and taking antigout medications. Patients not taking anti-gout medications and not showing unwillingness to participate were excluded. The eGFR was calculated with the CKD Epidemiology Collaboration (CKD-EPI) creatinine equation (2009). Doses to be individualized were decided based on the Renal Drug Handbook and BNF 80. Data were analyzed via R 4.0.3. Multinomial logistic regression analysis was performed to analyze statistical significance of risk with various predictors, considering a p-value <0.05 statistically significant. Comorbidities were coded as per the ICD-11 coding and ATC classification of medicines as per the WHO Guidelines for ATC classification and DDD assignment 2020. High risk of progression to CKD increased in the age range 54-63 and ≥84 years by 17.77 and 43.02 times, respectively. High risk of gout increased by 29.83 and 20.2 times for overweight and obese patients respectively. Aceclofenac 100 mg was prescribed for maximum patients (117, 30.5%). Need of dose individualization was realized altogether in case of 30 patients (8%), with maximum (7, 1.8%) in case of etoricoxib 90 mg (i.e., avoid if possible). There were 260 (67.7%) cases without associated comorbidities, followed by 37 (9.6%) patients with hyperthyroidism. Various glucocorticoids were prescribed for 141 (36.9%) patients, out of whom 14 (3.8%) required dose individualization. Altogether 61 (15.9%) patients were prescribed with xanthine oxidase inhibitors, out of whom 5 (1.3%) required dose individualization.

**Conclusions:** Thirty (8%) cases required dose individualization, which was although minimal but could have meaningful impact on clinical success of individual patient. Based on the recommendation on dose individualization, those patients could be optimized on their therapy on future follow ups. Also, future randomized controlled trials may be based on these to derive a more conclusive evidence base for gout management.

**Author summary:** *Why was this study done?:*

- Dosage individualization helps optimize drug selection and dosing based on pathogenesis, mechanism of action of drugs, and their dose exposure-response relationships.
- Very limited researches have been undertaken in Nepal to individualize medications for chronic medication users including gout patients. The present study was probably the first of its kind in Nepal aiming to individualize the dose of anti-gout medications among gout patients.

*What did the researchers do and find?:*

- Bicentric cross-sectional study was carried out among 384 new gout patients visiting two gout treatment centers in Nepal and taking antigout medications.
- Doses to be individualized were decided based on standard references of the Renal Drug Handbook and the British National Formulary 80. Dose to be individualized was later discussed with the prescribing physicians.
- Need of dose individualization was realized altogether in case of 30 patients (8%), with maximum (7, 1.8%) in case of etoricoxib 90 mg (i.e., avoid if possible).

*What do these findings mean?:*

- The present simple effort of individualization of antigout medications might help the physicians optimize the therapeutic success for the patients, once they implement it and patients adhere to it.
- Since the study was limited to only two gout and rheumatic diseases treatment centers, it might not represent the whole gout patients in the nation but it might provide the evidence base for future large controlled clinical trials.

## Introduction

Gout is an inflammatory arthropathy with a disorder of purine metabolism and is characterized by a sustained increase in blood uric acid, leading to urate crystal deposition and tissue damage [1,2]. Clinically, it manifests as the recurrent acute and chronic arthritis, tophi, urolithiasis and renal disease. Gouty arthritis and tophi may lead to chronic disability, impair health-related quality of life (HRQoL), increase healthcare resources and reduce productivity [2-4]. Gout has prevalence of 1.14%, 1.7%, 2.49%, 2.7% and 4% in China, Australia, UK, New Zealand, and US respectively [1,3,5,6] and its prevalence is rising every year both in developing and developed countries due to various factors such as high BMI (as obesity increases endogenous uric acid production), and concomitant diseases (e.g., hypertension), consumption of purine-rich diets (e.g., red meat, high-fructose syrup or beverage, beer, seafood), and medications (e.g., diuretics, cyclosporine) [4,6,7-10]. Country-wide gout prevalence is understudied domain in Nepal like other developing countries [11], and one research found a district-wise prevalence of 21.42% at Chitwan district, one of the districts out of 13 in Bagmati Province in Nepal [12].

Gout remains misunderstood, mis- or under-diagnosed, and undertreated despite various recommendations and guidelines on its management [5,8,13,14]. Guidelines for gout management usually help physicians select the most effective treatments and educate patients to promote adherence [13].

Acute gouty arthritis can be treated with colchicine, non-steroidal anti-inflammatory drugs (NSAIDs) (e.g., indometacin, naproxen), or glucocorticoids (oral and intra-articular injection) [8,10,15]. Xanthine oxidase inhibitors (XOIs) (e.g., allopurinol, febuxostat) are the first-line urate lowering therapies (ULTs) to prevent recurrent gouty arthritis whereas the uricosurics are the second-line medicines for those who cannot tolerate or show contraindications to XOIs [8]. Prophylaxis with colchicine should be administered followed by initiation of ULT (allopurinol or febuxostat). Patient education on risk factors for hyperuricaemia and gout can improve therapeutic success as it is generally managed suboptimally [9]. Also, shared decision-making between the informed patients and practitioners improves its effective management [14].

The conventional one-size-fits-all approach has been criticized for almost all drugs used especially for chronic diseases, including gout. Therefore, individualization of drug therapy has been put forth to tackle the problem as it helps optimize drug selection and dosing based on pathogenesis, mechanism of action (MOA) of drugs, and their dose exposure-response relationships. Dose individualization also helps optimize the efficacy of medicine by promoting its benefits and minimizing toxicity on individual patient. Dose can be individualized by various approaches such as selecting a medicine based on effective serum concentrations, population pharmacokinetics, therapeutic drug monitoring (TDM), pharmacogenomics and other [16]. Very limited efforts have been undertaken in Nepal to individualize medications for chronic medication users. The present study was probably the first of its kind in Nepal aiming to individualize and optimize the dose of anti-gout medications among gout patients.

## Methods

### Study design, study area and study population

Bicentric cross-sectional study was carried out among new gout patients visiting two gout treatment centers (i.e., Gosainkunda Health Care Clinic, Satdobato and Aarogya Health Home, Jawalakhel) at Lalitpur Metropolitan City, Nepal and taking antigout medications. Every day 10 to 15 patients came to the fist clinic whereas 50 to 80 patients came to the latter for gout treatment during the study period from July to December 2019.

### Ethics approval

Approval for research was obtained from the administrative department of both clinics, and ethics approval received from Nobel College Institutional Review Committee (NIRC), Sinamangal, Kathmandu (Ref. No.: BPY IRC215/2019). The patients were verbally informed about the research objectives and written informed consents were obtained from them prior to the initiation of research. Their confidentiality was maintained throughout the research period (i.e., pre- and post-completion of research).

### Sampling and sample size

Simple random sampling was followed for the data collection and sample size was calculated based on the formula:

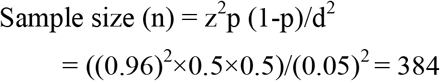

where, n = sample size; z = standard normal variate (1.96); p = population proportion; q = 1-p; d = absolute error or precision (0.05)

### Inclusion and exclusion criteria

All patients taking anti-gout medications along with laboratory reports consisting of creatinine value were included for the research. Patients not taking anti-gout medications and not showing unwillingness to participate in the research were excluded.

### Dosage individualization and optimization approaches

- Relevant patient-specific information such as demographic characteristics (e.g., age, gender, weight, height), comorbidities, diet restrictions, and gout medications related information were obtained from the patients’ prescriptions once they finished their physicians meeting.
- The estimated GFR (eGFR) was calculated with the Chronic Kidney Disease Epidemiology Collaboration (CKD-EPI) creatinine equation (2009) with the help of the National Kidney Foundation (NKF) GFR Calculator. The CKD-EPI creatinine equation was as follows: [17]

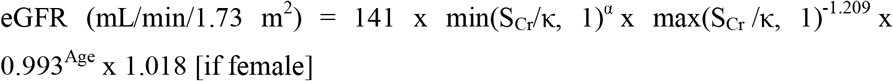

where, S_Cr_: standardized serum creatinine (mg/dL); κ = 0.7 (females) or 0.9 (males); α = -0.329 (females) or -0.411 (males); min = indicates the minimum of S_Cr_/κ or 1; max = indicates the maximum of S_Cr_/κ or 1

- The GFR categories and albuminuria categories were formed based on the criteria set by the NKF [17].
- Obesity status from the body mass index (BMI) was computed based on the CDC guidance [18].
- Finally, doses to be individualized were decided based on standard references of the Renal Drug Handbook 4^th^ edition [19] and the British National Formulary 80 [20]. Dose to be individualized was later discussed with the prescribing physicians.

### Data collection instrument, reliability and validity

We collected data in the self-developed pre-piloted questionnaire, developed based on the extensive literature review. Reliabilities of the tool developed and data collected on these were checked by Chronbach’s alpha technique, considering the benchmark of 70% for satisfactory level. Face validity, and content validity were checked based on the expert reviews and opinions on the questionnaire from five experts from the Department of Pharmaceutical Sciences, two experts from the Department of Medical Microbiology and one expert from the Department of Medical Laboratory Technology at Nobel College.

### Statistical analysis

We entered data into the IBM SPSS version 26 [21] and performed statistical testing via R 4.0.3 [22]. Categorical variables were presented with descriptive statistics. Multinomial logistic regression analysis was performed to analyze the statistical significance of risk of progression to chronic kidney disease (CKD) with various predictors (e.g., age, gender, obesity, eGFR, duration of disease, and diet restrictions), considering a p-value <0.05 statistically significant. Comorbidities of the patients were coded as per the International Statistical Classification of Diseases and Related Health Problems (ICD)-11 coding system [23]. The anatomic/therapeutic/chemical (ATC) classification of medicines was performed based on the WHO Guidelines for ATC classification and DDD assignment 2020 23rd edition [24].

## Results

There were 99 (25.8%) patients aged 44 - 53 years, 257 (66.9%) female, 194 (50.5%) participants with normal BMI (i.e., 18.5 to <25 kg/m2), 271 patients (70.6%) with eGFR 76 - 125 mL/min/1.73m2, 205 patients (53.4%) with normal or high GFR category (i.e., G1), 304 patients (79.2%) with duration of gout 0.2 - 10.1 years, and 260 patients (67.7%) on restriction of red meat, fish, alcohol and smoking, sour and spicy food, suffering from low risk of progression of CKD. (Table 1)

**Table 1:** Distribution of risk of progression of CKD with demographic and gout related characteristics of study population (n = 384)

| Study variables | Risk of progression of CKD |  |  | Total<br>(n,%) |
| --- | --- | --- | --- | --- |
|  | Low (n,%) | Moderate<br>(n,%) | Very high<br>(n,%) |  |
| Age (in years) (Mean±SD: 49.36±13.56) |  |  |  |  |
| <= 3 | 0 | 1 (0.3) | 0 | 1 (0.3) |
| 4 - 13 | 1 (0.3) | 0 | 0 | 1 (0.3) |
| 14 - 23 | 10 (2.6) | 0 | 0 | 10 (2.6) |
| 24 - 33 | 31 (8.1) | 0 | 0 | 31 (8.1) |
| 34 - 43 | 84 (21.9) | 6 (1.6) | 0 | 90 (23.4) |
| 44 - 53 | 99 (25.8) | 6 (1.6) | 0 | 105 (27.3) |
| 54 - 63 | 83 (21.6) | 7 (1.8) | 1 (0.3) | 91 (23.7) |
| 64 - 73 | 32 (8.3) | 6 (1.6) | 0 | 38 (9.9) |
| 74 - 83 | 10 (2.6) | 5 (1.3) | 0 | 15 (3.9) |
| 84+ | 1 (0.3) | 0 | 1 (0.3) | 2 (0.5) |
| Gender: |  |  |  |  |
| Male | 94 (24.5) | 6 (1.6) | 1 (0.3) | 101 (26.3) |
| Female | 257 (66.9) | 25 (6.5) | 1 (0.3) | 283 (73.7) |
| Obesity: |  |  |  |  |
| Underweight: BMI <18.5 kg/m2 | 31 (8.1) | 4 (1) | 0 | 35 (9.1) |
| Normal: BMI 18.5 to <25 kg/m2 | 194 (50.5) | 17 (4.4) | 0 | 211 (54.9) |
| Overweight: BMI 25 to <30 kg/m<sup>2</sup> | 106 (27.6) | 7 (1.8) | 1 (0.3) | 114 (29.7) |
| Obese: BMI ≥30 kg/m<sup>2</sup> | 20 (5.2) | 3 (0.8) | 1 (0.3) | 24 (6.3) |
| **eGFR with the CKD-EPI creatinine equation (2009) (mL/min/1.73m<sup>2</sup>)** (Mean±SD: 89.60±22.56) |  |  |  |  |
| ≤ 25 | 0 | 0 | 1 (0.3) | 1 (0.3) |
| 26 - 75 | 67 (17.4) | 31 (8.1) | 1 (0.3) | 99 (25.8) |
| 76 - 125 | 271 (70.6) | 0 | 0 | 271 (70.6) |
| 126 - 175 | 12 (3.1) | 0 | 0 | 12 (3.1) |
| 176+ | 1 (0.3) | 0 | 0 | 1 (0.3) |
| **GFR category & CKD classification:** |  |  |  |  |
| G1: GFR ≥90 mL/min/1.73 m<sup>2</sup> (Normal or high) | 205 (53.4) | 0 | 0 | 205 (53.4) |
| G2: GFR 60-89 mL/min/1.73 m<sup>2</sup> (Mildly decreased) | 146 (38) | 0 | 0 | 146 (38) |
| G3a: GFR 45-59 mL/min/1.73 m<sup>2</sup> (Mildly to moderately decreased) | 0 | 23 (6) | 0 | 23 (6) |
| G3b: GFR 30-44 mL/min/1.73 m<sup>2</sup> (Moderately to severely decreased) | 0 | 8 (2.1) | 0 | 8 (2.1) |
| G4: GFR 15-29 mL/min/1.73 m<sup>2</sup> (Severely decreased) | 0 | 0 | 2 (0.5) | 2 (0.5) |
| **Duration of disease** (Mean±SD: 4.92±5.27) |  |  |  |  |
| ≤ 0.1 | 7 (1.8) | 0 | 0 | 7 (1.8) |
| 0.2 - 10.1 | 304 (79.2) | 30 (7.8) | 2 (0.5) | 336 (87.5) |
| 10.2 - 20.1 | 35 (9.1) | 0 | 0 | 35 (9.1) |
| 20.2 - 30.1 | 3 (0.8) | 1 (0.3) | 0 | 4 (1) |
| 30.2+ | 2 (0.5) | 0 | 0 | 2 (0.5) |
| **Diet restriction:** |  |  |  |  |
| Beans, grains, sour and spicy foods, old pickle | 7 (1.8) | 0 | 0 | 7 (1.8) |
| Red meat, fish, alcohol and smoking, sour and spicy food | 260 (67.7) | 23 (6) | 2 (0.5) | 285 (74.2) |
| No restriction | 84 (21.9) | 8 (2.1) | 0 | 92 (24) |
| eGFR: Estimated glomerular filtration rate; GFR: Glomerular filtration rate; MDRD: Modification of Diet in Renal Disease |  |  |  |  |

The multinomial logistic regression analysis showed that moderate risk of progression to CKD decreased with advancing age but that of high risk increased in the age range 54-63 and ≥84 years by 17.77 and 43.02 times, respectively. The odds of male suffering from moderate and severe risks were 1/1.54 and 1/2.29e+07 respectively, showing less risk of gout among males compared to females. High risk of gout increased by 29.83 and 20.2 times for overweight and obese patients respectively. High risk of CKD progression decreased with all eGFR values from 26 to ≥176 whereas the moderate risk decreased for eGFR from 126 to ≥176. Moderate risk mainly increased by 20.47 and 34.29, provided that the duration of gout was 0.2-10.1 and 20-30.1 years. (Table 2)

**Table 2:**
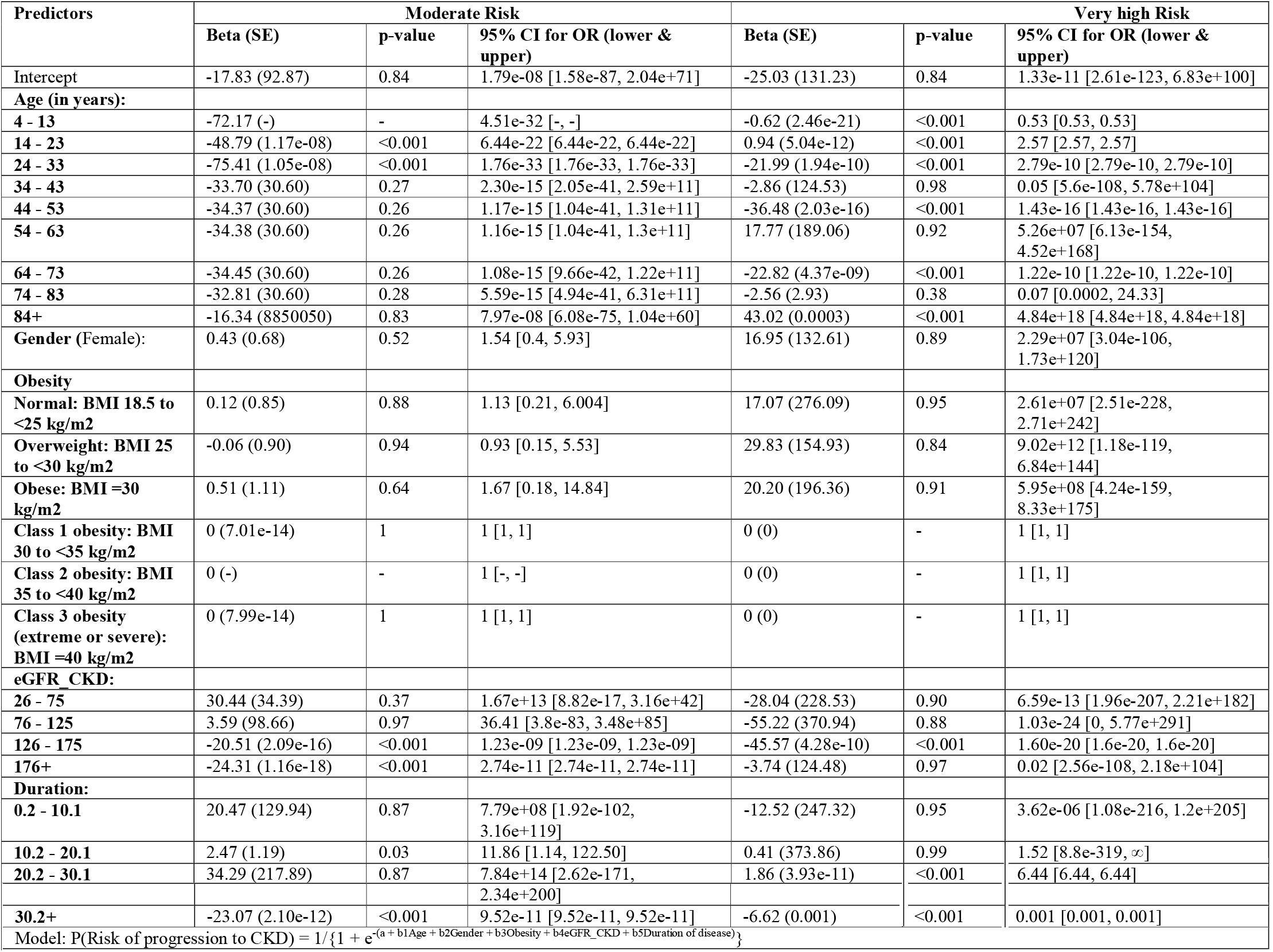
Multinomial regression of risk of progression to CKD (with reference to low risk) with various predictors.

Aceclofenac 100 mg was prescribed for maximum patients (117, 30.5%). Need of dose individualization was realized altogether in case of 30 patients (8%), with maximum (7, 1.8%) in case of etoricoxib 90 mg (i.e., avoid if possible). (Table 3) There were 260 (67.7%) cases without associated comorbidities, followed by 37 (9.6%) patients with hyperthyroidism. (Annex 1) Altogether 220 (57.5%) patients were prescribed with various NSAIDs, for whom 7 (1.8%) patients required dose individualization, all for etoricoxib 90 mg. Various intermediate-acting glucocorticoids were prescribed for 141 (36.9%) patients, out of whom 14 (3.8%) required dose individualization. Altogether 61 (15.9%) patients were prescribed with XOIs, out of whom 5 (1.3%) required dose individualization. (Annex 2)

**Table 3:** Medicines used with the status of dose individualization (n = 384)

| SN | Medicines | Therapeutic category | ATC Classification* | Not used (n,%) | Used (n,%) | No Dose adjustment required (n,%)** | Dose individualization required (n,%) (Total: 30, 8%)** | Individualized dose |
| --- | --- | --- | --- | --- | --- | --- | --- | --- |
| 1 | Aceclofenac 100 mg | Preferential COX-2 inhibitor | M01A | 267 (69.5) | 117 (30.5) | 117 (30.5) | - | NA |
| 2 | Allopurinol 100 mg | XOI | M04AA | 383 (99.7) | 1 (0.3) | 1 (0.3) | - | NA |
| 3 | Colchicine 500 mcg BD | Alkaloid | M04AC01 | 344 (89.6) | 40 (10.4) | 36 (9.4) | Reduce dose or dosage interval by 50%: 4 (1) | 250 mcg |
| 4 | Etoricoxib 60 mg | NSAIDs: COX-2 inhibitor | M01AH05 | 383 (99.7) | 1 (0.3) | 1 (0.3) | - | NA |
| 5 | Etoricoxib 90 mg |  |  | 292 (76) | 92 (24) | 85 (22.1) | Dose as in normal renal function, but avoid if possible: 7 (1.8) | Avoid if possible |
| 6 | Etoricoxib 90/60 mg |  |  | 383 (99.7) | 1 (0.3) | 1 (0.3) | - | NA |
| 7 | Febuxostat 40 mg | XOI | M04AA | 331 (86.2) | 53 (13.8) | 49 (12.8) | Start with 40 mg and monitor closely: 1 (0.3);<br>Dose as in normal renal function: 3(0.8) | 40 mg with close monitoring |
| 8 | Febuxostat 80 mg |  |  | 377 (98.2) | 7 (1.8) | 6 (1.6) | Dose as in normal renal function: 1(0.3) | Dose as in normal renal function |
| 9 | Indometacin 25 mg | NSAIDs: Indole acetic acid derivative | M01AB01 | 383 (99.7) | 1 (0.3) | 1 (0.3) | - | NA |
| 10 | Indometacin 50 mg |  |  | 378 (98.4) | 6 (1.6) | 6 (1.6) | - | NA |
| 11 | Methylprednisolone inj. 80 mg | Glucocorticoid: Intermediate acting | H02AB04 | 355 (92.4) | 29 (7.6) | 28 (7.3) | Dose as in normal renal function: 1(0.3) | Dose as in normal renal function |
| 12 | Naproxen 250 mg | NSAIDs: Propionic acid derivatives | M01AE02 | 382 (99.5) | 2 (0.5) | 2 (0.5) | - | NA |
| 13 | Prednisolone 2.5 mg | Glucocorticoid: Intermediate acting | H02AB06 | 352 (91.7) | 32 (8.3) | 29 (7.6) | Dose as in normal renal function: 3(0.8) | Dose as in normal renal function |
| 14 | Prednisolone 5 mg |  |  | 354 (92.2) | 30 (7.8) | 27 (7) | Dose as in normal renal function: 3(0.8) | Dose as in normal renal function |
| 15 | Prednisolone 10 mg |  |  | 372 (96.9) | 12 (3.1) | 10 (2.6) | Dose as in normal renal function: 2(0.5) |  |
| 16 | Prednisolone 15 mg |  |  | 379 (98.7) | 5 (1.3) | 3 (0.8) | Dose as in normal renal function: 2(0.5) |  |
| 17 | Prednisolone 20 mg |  |  | 374 (97.4) | 10 (2.6) | 9 (2.3) | Dose as in normal renal function: 1(0.3) |  |
| 18 | Prednisolone 25 mg |  |  | 382 (99.5) | 2 (0.5) | 2 (0.5) | - | NA |
| 19 | Prednisolone 30 mg |  |  | 369 (96.1) | 15 (3.9) | 14 (3.6) | Dose as in normal renal function: 1(0.3) | Dose as in normal renal function |
| 20 | Prednisolone 30/20/10 |  |  | 383 (99.7) | 1 (0.3) | 1 (0.3) | - | NA |
| 21 | Prednisolone 20/15/10 |  |  | 383 (99.7) | 1 (0.3) | 1 (0.3) | - | NA |
| 22 | Prednisolone 15/10/5/2.5 mg |  |  | 383 (99.7) | 1 (0.3) | 1 (0.3) | - | NA |
| 23 | Prednisolone 15/10/5 |  |  | 383 (99.7) | 1 (0.3) | 1 (0.3) | - | NA |
| 24 | Prednisolone 5/2.5 |  |  | 383 (99.7) | 1 (0.3) | - | Dose as in normal renal function: 1(0.3) | Dose as in normal renal function |
| 25 | Tapentadol 50 mg | Atypical opioid | D06007 | 381 (99.2) | 3 (0.8) | 3 (0.8) | - | NA |
| 26 | Triamcinolone 10 mg inj. | Glucocorticoid: Intermediate acting | D07XB02 | 383 (99.7) | 1 (0.3) | 1 (0.3) | - | NA |

NA: not applicable (i.e., as usual dose )
\*WHO Guidelines for ATC classification and DDD assignment 2020
\*\* The Renal Drug Handbook: The ultimate prescribing guide for renal practitioners 4th edition (2014)

## Discussion

Maximum patients (i.e., 266, 69.3%) aged 34 to 63 years were suffering from low risk of progression to CKD with gout, and the high risk increased in the age range 54-63 and ≥84 years by 17 and 43 times, respectively. The finding was in line with various published findings that gout mainly affects men over the age of 40 years [6,7,14,25-27]. Such slight variation might be due to the inclusion of all age groups in the present research. Since glomerular filtration rate (GFR) is regarded as the best index of kidney function based on the data of serum creatinine, age, gender, race and/or body weight and usually declines with advanced age, gout may be more common phenomenon among the aged [17].

Maximum female (257, 66.9%) patients were suffering from low risk of gout during the study period. However, various researches found that gout is three times more common among men compared to women [8,14], female accounting for only 5% of all gout patients, although in increasing trend [5,10]. The incongruence in case of gender might be the reflection of the participation of more number of female patients within the study period. Also, deposition of monosodium urate crystals in any of the age groups in gender non-specific pattern may lead to chronic arthritis, tophi, urolithiasis and renal disease [26]. Further, gout may be misdiagnosed due to insidious attack or diagnosis is usually delayed among women due to the protective effects of estrogen during premenopause and the incidence rising after menopause and peaking at ≥80 years among women [5,10].

Overweight men with BMI >27.5 kg/m2 usually have up to 16 times higher risk for gout than men with normal BMI with <20 kg/m2 [9]. The present research also showed that high risk of gout increased by 29 and 20 times for overweight and obese patients respectively. This might be aggravated by comorbidities as 260 (67.7%) cases without associated comorbidities, followed by 37 (9.6%) patients with hyperthyroidism seemed to have risk of gout. Other researches also concluded that gout may appear with various co-morbidities including obesity, dyslipidemia, cardiovascular disease (e.g., diabetes mellitus, hypertension), CKD, hypothyroidism, anemia, chronic pulmonary diseases [2-4].

The eGFR is a reliable diagnostic test to detect and manage kidney disease, plan for dialysis, evaluate the success of renal transplantation, if any, and to optimize medications dosing, and it gets declined with advancing age [17]. High risk of CKD progression decreased with all eGFR values from 26 to ≥176 whereas the moderate risk decreased with eGFR from 126 to ≥176. Moderate risk increased by 20.47 and 34.29 times, provided that the duration of gout was 0.2-10.1 and 20-30.1 years, indicating that the more chronic the disease becomes, the more deteriorating effects might appear.

The research found that 260 low risk patients (67.7%) were suggested to remain on restriction of red meat, fish, alcohol and smoking, sour and spicy food to prevent from their progression to CKD. However, Wise (2005) reported that only the dietary restrictions alone may not have major impacts on the elderly gout patients, and even purine-free diets may have low effect on maintenance of target serum urate [28]. However, treatment approaches are the same for both men and women with gout including avoidance of risk factors (e.g., alcohol, diuretics), maintenance of normal serum glucose levels, blood pressure, body weight and BMI, and less consumption of purine-rich diets (e.g., red meat, fructose, seafood) [10].

### Dose individualization approaches

Acute gout flare can be treated and even the prophylaxis can be provided with colchicine, NSAIDs, and corticosteroids [9] and 40 (10.4%), 220 (57.5%) and 141 (36.9%) patients were prescribed with these categories of medicines in the present research, either alone or combined. Four (1%) of colchicine users required dose adjustment. Colchicine can be used for management of both acute and chronic gout due to its anti-inflammatory effects. Its lower starting doses (i.e., maximum of 3 tablets of 500 mcg in the first 24 hour) are preferred to avoid untoward effects without compromising efficacy [29]. Colchicine 500 mcg twice daily, and 500 mcg/day (i.e., reduced frequency) can be used by the patients with normal renal function and renal impairment respectively [30], which was seemed to the case in the present research too. Renal function status should be assessed before initiating colchicine or NSAIDs. ([26]) Colchicine is avoided in the elderly as this is poorly tolerated [31]. Colchicine or NSAID prophylaxis should be given when antihyperuricemic therapy is initiated and should be continued for a prolonged period till the patient is free from gout attacks [32].

Seven (1.8%) of NSAIDs users required dose adjustment and such dose individualization was proposed for etoricoxib 90 mg (i.e., dose as in normal renal function, but avoid if possible). NSAIDs with short half-lives (<6 hr) (e.g., diclofenac and ketoprofen) generally reach steady-state levels more quickly than those with long half-lives (i.e., >6 hr) and are preferred in patients with acute gout with concurrent renal failure, uncontrolled hypertension or cardiac failure [29,31]. However, none of the patients in the present research were prescribed with these particular NSAIDs. NSAIDs should be initiated at its maximum dose, rather than titrating upwards from the lower dose in case of acute gout [14,29]. For e.g., indometacin 150-200 mg/day, naproxen 1000 mg/day, diclofenac 150 mg/day are used for 4-8 days and then gradually tapered till symptoms get resolved [29]. But the case was not evident in the present study as indometacin was found to be prescribed in 25 and 50 mg doses, and naproxen 250 mg dose, with no diclofenac being prescribed. Although NSAIDs are preferred to colchicine due to their fewer side effects [29], still, they are not risk-free, and their use is limited by the risks of gastropathy (e.g., gastric ulceration and gastritis), GI bleeding, acute renal failure, fluid retention, and reduced effectiveness of antihypertensives with their excessive consumption [26,28] Indometacin should be avoided and other NSAIDs should be used in low doses for short periods by the elderly with acute gout [28]. Also, oal NSAIDs are generally less cost-effective than oral corticosteroids or colchicine, due to their adverse effects [14]. NSAIDs are usually contraindicated in the elderly and low-dose colchicine (500 mcg/day) or prednisone 5–7.5 mg/day may be used for the first 6 months after initiation of allopurinol [33].

Fourteen (3.8%) of the intermediate-acting glucocorticoids users required dose individualization in the present study. Systemic corticosteroids can be used for treating acute gout when NSAIDs and colchicine are contraindicated [14,29]. Prednisone can be started at a dose of 20-40 mg/day and gradually tapered and discontinued over 10-14 days. However, corticosteroids should be cautiously used in case of diabetes mellitus, hypertension, congestive heart failure (CHF), coronary artery disease (CAD) and severe infections [29].

Sixty one (15.9%) patients were prescribed with xanthine oxidase inhibitors (XOIs), out of whom 5 (1.3%) required dose individualization, all for febuxostat users. The first-line ULT (i.e., allopurinol) was found to be prescribed for only a single patient, whereas the second-line ULT (i.e., febuxostat) for 60 patients (15.6%) as it is used if allopurinol is not tolerated or the response is inadequate ([1]; [2]) or when uricosurics are contraindicated (as in case of stage 3 CKD), urolithiasis, or uric acid overproduction [34]. Febuxostat might have been preferred to allopurinol in the present study due to its few drug interactions compared to the latter and its minimal propensity to cause allopurinol hypersensitivity syndrome (AHS) [30,35-37]. Also, it has added benefit that its dose adjustment is usually not necessary in case of mild to moderate renal or liver insufficiency or advanced age [8,37-39]. However, other second-line ULTs (i.e., probenecid, benzbromarone) were not prescribed for any of the patients enrolled in the study.

Febuxostat 40 mg was prescribed for 53 (13.8%) whereas its 80 mg dose for 7 (1.8%) patients in the present research. Majority of febuxostat users were started with a dose of 40 mg/day as expected but 80 mg could be used in case serum uric acid levels not achieving <6 mg/dL even two weeks after initiation of therapy [38]. It was proposed to start febuxostat with 40 mg with close monitoring in case of one (0.3%) patient, and dose as in normal renal function in case of 3 (0.8%) patients prescribed with 40 mg dose, and dose as in normal renal function in case of one (0.3%) patient prescribed with 80 mg dose. Febuxostat 40 mg/day has been found to be clinically equivalent in efficacy to allopurinol 300 mg/day ([40]; [30]; [39]) and its 80 mg/day dose has superior urate-lowering efficacy compared to allopurinol 300 mg/day [37,40]. Its dose can be increased to 120 mg/d after 4 weeks to reach the therapeutic target for sUA levels [26,38] but none of the patients reached up to that level in the present study.

Medications can be effective only when used at optimal doses and if individuals fully adhere to the regimens. However, adherence to ULTs for gout management is usually not up to the satisfactory level because of various reasons including the risk and persistence of acute gout flare even with their initiation, poor response to these, suboptimal dosing and intolerance to these. Poor adherence and non-adherence (either primary or secondary) to medications prescribed for the management of chronic diseases such as gout are a global problem as the persistent untreated gout can lead to recurrent gouty arthritis, chronic gouty arthropathy, tophi and urolithiasis [41]. Patients should be educated on lifestyle modifications (e.g., exercise, consumption of low purine diets) along with medications to reduce the raised serum urate levels. Weight reductions with exercise show a positive impact on urate reduction [32].

### Strengths and limitations of study

The report of individualization status based on the Renal Drug Handbook was provided to the concerned physicians so that they might be aware of the need of the same for the patients in their scheduled follow ups. This might help the physicians optimize the therapeutic success, once they implement it and patients adhere to it. However, the present study was not free from some of the limitations including:

- The study was limited to only two gout and rheumatic diseases treatment centers, which might not represent the whole gout patients in the nation.
- Large controlled clinical trials could be conducted based on the foundation of the present study.

## Conclusions

Majority of patients (220, 57.5%) were prescribed with NSAIDs and 7 (1.8%) of whom required dose individualization. Thirty (8%) cases required dose individualization, which was although minimal but could have meaningful impact on the clinical success of the individual patient. Based on the recommendation on the dose individualization of the present study, those patients could be optimized on their therapy on their future follow ups, providing optimal benefits to them. Also, future randomized controlled trials may be based on the findings of the present study to derive a more conclusive evidence base for gout management.

## Data Availability

All data supporting the findings of this study are contained within the manuscript. Any additional information regarding the study including the questionnaires would be shared by the corresponding author upon request.

## Acknowledgements

The authors are grateful to all the respondents for their valuable time and active cooperation throughout the research. They are also grateful to the physicians and the administrators at the two study clinics who provided their intellectual and managerial support throughout the research phase.

## Competing interests

The authors declare that they have no competing interests.

## Authors’ Contributions

BS conceptualized and designed the study, performed literature review, analyzed and interpreted data, and prepared the final manuscript. SC, PG, AH and SS designed the study, performed literature review, did necessary fieldworks including data collection, performed literature review, drafted and revised the manuscript. All authors read and approved the final manuscript.

## Declarations

### Ethics approval and consent to participate

The study was ethically approved by the Nobel College Institutional Review Committee, Sinamangal, Kathmandu (Ref. No.: BPY IRC215/2019).

### Consent for publication

Not applicable

### Funding

None

### Conflicts of interest

The authors declare no potential conflicts of interest related to the present research and publication.

## Notes

### Competing Interest Statement

The authors have declared no competing interest.

### Author Declarations

Approval for research was obtained from the administrative department of both clinics, and ethics approval received from Nobel College Institutional Review Committee (NIRC), Sinamangal, Kathmandu (Ref. No.: BPY IRC215/2019).

