## Supplemental Table 1 and 2 for "Dosage individualization proposed for anti-gout medications among the patients with gout: a bicentric study in Nepal"

**S1 Appendix: Comorbidities of the patients** (WHO, 2020)

| **Comorbid conditions** | **ICD-11 classification*** | **Frequency (%)** |
| --- | --- | --- |
| Allergic reactions | 4A8Z | 1 (0.3) |
| Asthma | CA23 | 2 (0.5) |
| DM-2 | 5A11 | 7 (1.8) |
| Gastritis | DA42 | 1 (0.3) |
| Hyperlipidemia | 5C80.Z | 1 (0.3) |
| Hypertension | BA00 | 30 (7.8) |
| Hypertension, DM-2 | BA00, 5A11 | 9 (2.3) |
| Hypertension, DM-2, Hyperlipidemia | BA00, 5A11, 5C80.Z | 9 (2.3) |
| Hypertension, DM-2, Hyperthyroidism | BA00, 5A11, 5A02.Z | 3 (0.8) |
| Hypertension, Gastritis | BA00, DA42 | 1 (0.3) |
| Hypertension, Hyperthyroidism | BA00, 5A02.Z | 1 (0.3) |
| Hypertension, Renal impairment | BA00, GB6Z | 2 (0.5) |
| Hyperthyroidism | 5A02.Z | 37 (9.6) |
| Hyperthyroidism, DM-2 | 5A02.Z, 5A11 | 2 (0.5) |
| Hyperthyroidism, Hyperlipidemia | 5A02.Z, 5C80.Z | 1 (0.3) |
| Hyperthyroidism, Vitamin deficency | 5A02.Z, 5B55 | 1 (0.3) |
| Hypotension | BA20 | 2 (0.5) |
| Migraine | 8A80 | 11 (2.9) |
| No comorbid condition | QA02 | 260 (67.7) |
| Psychiatric disorder, Hyperthyroidism | 6E8Z, 5A02.Z | 1 (0.3) |
| Renal impairment | GB6Z | 2 (0.5) |
| Total |  | 384 (100) |
| ICD: International Statistical Classification of Diseases and Related Health Problems | | |

**S2 Appendix: Therapeutic category-wise medication use**

| **SN** | **Therapeutic category** | **Medicines** | **Used** | **No dose adjustment required (n,%)** | **Dose adjustment required** |
| --- | --- | --- | --- | --- | --- |
| 1 | NSAIDs | Aceclofenac 100 mg | 117 (30.5) | 117 (30.5) | - |
|  |  | Etoricoxib 60 mg | 1 (0.3) | 1 (0.3) | - |
|  |  | Etoricoxib 90 mg | 92 (24) | 85 (22.1) | 7 (1.8) |
|  |  | Etoricoxib 90/60 mg | 1 (0.3) | 1 (0.3) | - |
|  |  | Indometacin 25 mg | 1 (0.3) | 1 (0.3) | - |
|  |  | Indometacin 50 mg | 6 (1.6) | 6 (1.6) | - |
|  |  | Naproxen 250 mg | 2 (0.5) | 2 (0.5) | - |
|  | Total |  | 220 (57.5) | 213 (55.6) | 7 (1.8) |
| 2 | XOIs | Allopurinol 100 mg | 1 (0.3) | 1 (0.3) | - |
|  |  | Febuxostat 40 mg | 53 (13.8) | 49 (12.8) | 4 (1) |
|  |  | Febuxostat 80 mg | 7 (1.8) | 6 (1.6) | 1(0.3) |
|  | Total |  | 61 (15.9) | 56 (14.7) | 5 (1.3) |
| 3 | Alkaloids | Colchicine 500 mcg BD | 40 (10.4) | 36 (9.4) | 4 (1) |
| 4 | Glucocorticoids (Intermediate acting) | Methylprednisolone inj. 80 mg | 29 (7.6) | 28 (7.3) | 1(0.3) |
|  |  | Prednisolone 2.5 mg | 32 (8.3) | 29 (7.6) | 3(0.8) |
|  |  | Prednisolone 5 mg | 30 (7.8) | 27 (7) | 3(0.8) |
|  |  | Prednisolone 10 mg | 12 (3.1) | 10 (2.6) | 2(0.5) |
|  |  | Prednisolone 15 mg | 5 (1.3) | 3 (0.8) | 2(0.5) |
|  |  | Prednisolone 20 mg | 10 (2.6) | 9 (2.3) | 1(0.3) |
|  |  | Prednisolone 25 mg | 2 (0.5) | 2 (0.5) | - |
|  |  | Prednisolone 30 mg | 15 (3.9) | 14 (3.6) | 1(0.3) |
|  |  | Prednisolone 30/20/10 | 1 (0.3) | 1 (0.3) | - |
|  |  | Prednisolone 20/15/10 | 1 (0.3) | 1 (0.3) | - |
|  |  | Prednisolone 15/10/5/2.5 mg | 1 (0.3) | 1 (0.3) | - |
|  |  | Prednisolone 15/10/5 | 1 (0.3) | 1 (0.3) | - |
|  |  | Prednisolone 5/2.5 | 1 (0.3) | - | 1(0.3) |
|  |  | Triamcinolone 10 mg inj. | 1 (0.3) | 1 (0.3) | - |
|  | Total |  | 141 (36.9) | 127 (33.2) | 14 (3.8) |
| 5 | Atypical opioid | Tapentadol 50 mg | 3 (0.8) | 3 (0.8) | - |
| DHFRase: Dihydrofolate reductase; SYSADOAs: symptomatic slow-acting drug in osteoarthritis; XOI: Xanthine oxidase inhibitor | | | | | |
